# The Integrated Voxel Analysis Method (IVAM) to Diagnose Onset of Alzheimer’s Disease and Identify Brain Regions through Structural MRI Images

**DOI:** 10.1101/19009597

**Authors:** Matthew Hur, Armen Aghajanyan

## Abstract

Magnetic Resonance Imaging (MRI) provides three-dimensional anatomical and physiological details of the human brain. We describe the Integrated Voxel Analysis Method (IVAM) which, through machine learning, classifies MRI images of brains afflicted with early Alzheimer’s Disease (AD). This fully automatic method uses an extra trees regressor model in which the feature vector input contains the intensities of voxels, whereby the effect of AD on a single voxel can be predicted. The resulting tree predicts based on the following two steps: a K-nearest neighbor (KNN) algorithm based on Euclidean distance with the feature vector to classify whole images based on their distribution of affected voxels and a voxel-by-voxel classification by the tree of every voxel in the image. An Ising model filter follows voxel-by-voxel tree-classification to remove artifacts and to facilitate clustering of classification results which identify significant voxel clusters affected by AD. We apply this method to T1-weighted MRI images obtained from the Open Access Series of Imaging Studies (OASIS) using images belonging to normal and early AD-afflicted individuals associated with a Client Dementia Rating (CDR) which we use as the target in the supervised learning. Furthermore, statistical analysis using a pre-labeled brain atlas automatically identifies significantly affected brain regions. While achieving 90% AD classification accuracy on 198 images in the OASIS dataset, the method reveals morphological differences caused by the onset of AD.

## Introduction

Alzheimer’s Disease (AD) is a prevalent degenerative disorder in today’s society as the 7th leading cause of death in America (Speert et al., 2012). As the causes and inner mechanisms underlying AD-related brain abnormalities are not fully understood, no cure has yet been found; however, treatments such as pharmacology that inhibit acetylcholinerase have successfully prolonged the lifespan of affected individuals by slowing down the degeneration of acetylcholine-releasing neurons (Bianchetti et al., 2006). Other biomolecular phenomena including the formation of beta-amyloid plaques and tao-fibrillary tangles have been implicated, but its relation to macroscopic mechanisms concerning brain regions is still nebulous. Computational methods for classification and segmentation can facilitate and supplement clinical diagnoses.

In order to reveal these macroscopic mechanisms between brain regions, many current image classification and segmentation algorithms incorporate an essential step of feature extraction. For example, the widely used and effective Voxel-Based Morphometry (VBM) method is designed for feature extraction to determine specific anatomic patterns of cerebral atrophy (Ashburner and Friston, 2000). However, this method suffers from its dependence on a precise registration and warping of MRI images to *a priori* probability maps (Veress et al., 2013). Its high sensitivity to accurate registration creates a limitation because the templates inherently differ with various MRI images due to structural variance of brain shape. Additionally, its high computational complexity leads to complex implementation and long run-time. Other methods for segmentation such as the Hybrid Watershed Algorithm (HWA) and the skull-stripping Brain Extraction Methods (BEM and BEM2DE) rely on accurate iterative thresholding and on assumptions about brain shape that limit their practical use when analyzing variegated brain shapes belonging to subjects of various demographic groups (Fennema-Notestine et al., 2006). Fully automatic methods need to quickly and effectively account for individual differences in brain shape without human supervision.

The essential steps that improve the results in these automatic methods are energy-based deformation fields which identify regions of interest, whereby a driving force pushes an objective function to convergence. Success of the energy methods can be attributed to the utilization of information about local differences as well as about global trends of the image.

We propose a multifaceted algorithm that utilizes methods for decision-tree learning to robustly and automatically classify AD affected brains as well as cluster and segment classification results of individual voxels to yield severely affected brain regions (Figure 1). First, the method, termed the Integrated Voxel Analysis Method (IVAM), skull-strips each test and training MRI image and spatially normalizes to the MNI152 brain-masked, i.e. skull-stripped, template. At the center of our framework of integrated processes is a trained extra-randomized trees regressor known to be highly effective for supervised learning of complex data (Geurts et al., 2006). The trained model is used to estimate the distribution of Alzheimer affected voxels, which are sampled from the MRI. A second model placed on top of the distributions predicts the CDR of the whole brain. The aggregated learning model can also automatically relay information about specifically affected brain regions allowing for novel insights into the inner neurological workings of AD tailored to accurately diagnose AD in its early stages.

**Figure 1:**
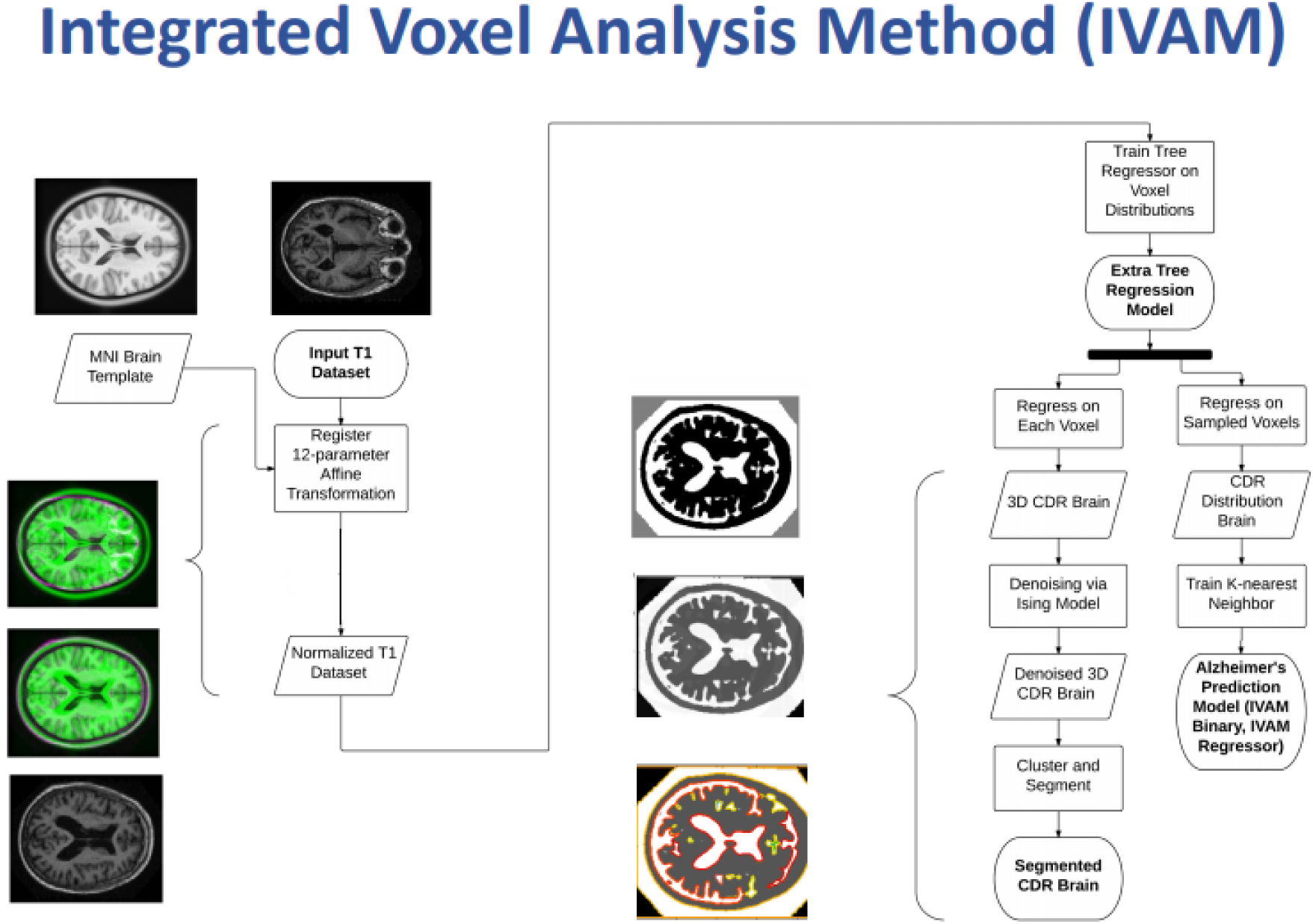
A flowchart outlining the Integrated Voxel Analysis Method (IVAM) with cross-sections resembling certain steps. The preprocessing step incorporates spatial normalization and (not shown) skull-stripping to create a dataset applicable to sampling for IVAM. The machine learning section utilizes a K-Nearest Neighbor (KNN) classifier which takes as input the voxel-by-voxel classified brain from an extra-randomized regressor tree.

## Methods

### A. Data Set

We obtain 233 anonymous MRI images from the Open Access Series of Imaging Studies (OASIS) (Buckner et al., 2004). Their age range is from 18-96 years with a mean of 53 years with each clinically diagnosed with a Client Dementia Rating (CDR) which rates subjects based on 6 criteria: memory, orientation, judgment and problem solving, function in the community, home and hobbies, and personal care (Buckner et al., 2004). The values range from 0 to 3 where 0 indicates no dementia while 0.5 and 1 indicate very mild and mild dementia respectively. In order to specifically analyze the onset of AD and due to the sparsity of the dataset of images labeled with a CDR > 1, we use images labeled with a CDR of 0, 0.5, and 1. All images are T1-weighted prepared rapid echo-gradient and obtained on a 1.5-T vision scanner by the OASIS study. (Buckner et al., 2004). Acquisition matrix was [256 256] with 128 contiguous saggital slices of thickness of 1.25 mm.

### B. Skull-Stripping

We perform two gray-level thresholds on the raw MRI images using Matlab. Then, we dilate the image with a ball structuring image, perform a third gray-level threshold, and isolate the main part of the head by taking the largest connected component in 3D.

### C. Spatial Normalization

One of the most essential components to accurate analysis of MRI images is the spatial normalization of the dataset. Through co-registration, processing of MRI images can be compared across multiple subjects, especially important for brain Region of Interest (ROI) analysis in pinpointing the affected regions. The images were normalized to the template provided by the Montreal Neuroimaging Institute known as the MNI152 template built by averaging across 152 brains. The images were first converted to the same voxel resolution of 1mm × 1mm × 1mm and symmetrically zero-padded to transform to the same dimensions. Next, each image was registered to the MNI152 template using a one plus one evolutionary optimizer to maximize the mutual information metric provided by David Mattes (Mattes et al., 2001).

### D. Feature Selection

Feature selection algorithm relied on the assumption that the level of Alzheimer’s projected at a voxel depends on the surrounding voxels as well as the three-dimensional coordinate of the voxel. Our feature vector included the values of all voxels around the voxel of interest with a preset radius and the appended 3D coordinates of the voxels of interest as well as the gender of the subject as male and female brain anatomy have been shown to exhibit structural differences (Ritchie *et al*. 2018). For each MRI image we enumerate all coordinates where the surrounding points are not all zeros. For each voxel we selected features as described above. Because of the sheer size of MRI images, we added a variable step constant throughout the whole aggregated model. Instead of enumerating every point, the step dictates step incrementation in the coordinates for the voxel of interest.

### E. Building the Model

The label of each feature vector is simply the CDR rating of respective patient. Once we built the data-set, we randomly sampled half of the data and used that to train a the ensemble, Extra Random Trees Regressor (Geurts et al., 2006) using the popular sklearn library (Pedregosa et al., 2011). Although traditionally combined with an ensemble method such as Random Forests, these Extra Random Tree Regressors (ERTR) tend to exhibit losses in accuracy observed through computational trials we conducted compared to the ERTR alone. The ERTR now predicts the CDR rating of single voxels in the MRI image.

### F. Predicting the CDR

Our method for predicting the CDR arises from our axiomatic assumption that the severity of Alzheimer’s disease in a patient, will be represented by the distribution of individual CDR regressed voxels. Our first intuition was to use the Kullback-Leibler divergence as a tool to compare the distributions. Although this gave us promising results, the utilization of K-Nearest Neighbor (Bentley, 1975) achieved much greater accuracy than the Kullback-Leibler divergence. Voxel sampling from the brain proceeded in the same way as feature selection, using the same function. After sampling, the regression model predicted the CDR of each individual voxel. The label or target for the KNN algorithm arises as a function of the CDR of the respective patient. The accuracy will be introduced at the Discussion section below.

### G. Identifying Severely Affected Brain Regions

The process of pinpointing severely affected brain regions from MRI’s can be seen as a pipelining process. A sagittal half-brain slice of a normalized MRI image compares to the following CDR classified image: the classified image depicts the results of the denoising model applied to the CDR voxel predictions. Black, grey, and white correspond to CDR predictions of 0, 0.5, and 1 respectively.

### H. Denoising via Ising Model

Using Murphy’s derivation of the Ising model, we define the probability of the update as

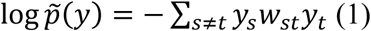

(Murphy, 2012).

The weight wst signifies the amount we attribute to the difference between the two pixels from each 2D slice and y_*n*_ represents the intensity value of voxel y. To simplify the calculations, we modify our equation to:

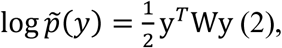

where W is a Toeplitz matrix. We define our objective function as

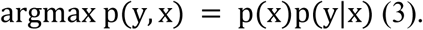

Continuing, by Murphy’s derivation, the unnormalized prior develops as follows:

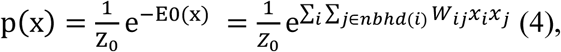

where nbhd denotes the immediate neighborhood of the two-dimensional pixel j. The Z_0_ is not needed since we are using the unnormalized prior. On the other hand, our unnormalized posterior will be:

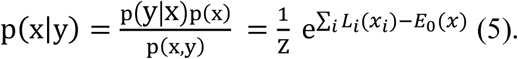

For the mean field update, we need to compute (see Murphy section 21.3.1 for details)

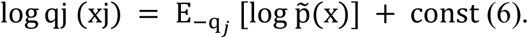

and since

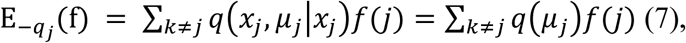

we have

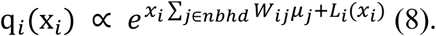

which yields the important theoretical step. Murphy derives an actual update. In Murphy’s (Murphy, 2012) derivation (page 738), note that it uses

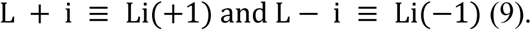

which are the log likelihood functions centered at each of these two values. The variance in the likelihood controls the strength of the prior. This is the final update, which also incorporates a damping term:

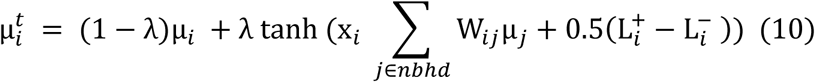

To convert this algorithm into a tertiary denoiser, we introduced this function to convert CDR to Ising denoiser values,

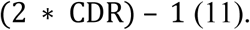

### I. Clustering and Segmentation Model

Our clustering and segmentation model bases on Ward’s method (Ward, 1963). We input the connectivity matrix of the denoised image into Ward’s hierarchical clustering method. The connectivity matrix can be defined as a matrix where each sample is defined through the neighboring samples following a given structure of the data. The connectedness structure of the data consisted of the one-voxel neighborhood of each voxel.

## Results

The hippocampus, heavily involved in memory storage, appears highly affected at both 0.5 and 1.0 CDR according to IVAM. In a sagittal cross-section, the streak above the hippocampus known as the inferior sagittal sinus appears slightly less but still highly affected by early Alzheimer’s Disease.

To automate the finding of affected brain regions, we downloaded the Talairach labeled database and registered it to the MNI template, then through a series of 90 degree 3D rotations, matched it to the orientation of 3D classified brains. Testing the difference between brain regions of CDRs of 0 (n = 51) and 0.5 (n = 11), the results are shown in Table 1.

**Table 1:**
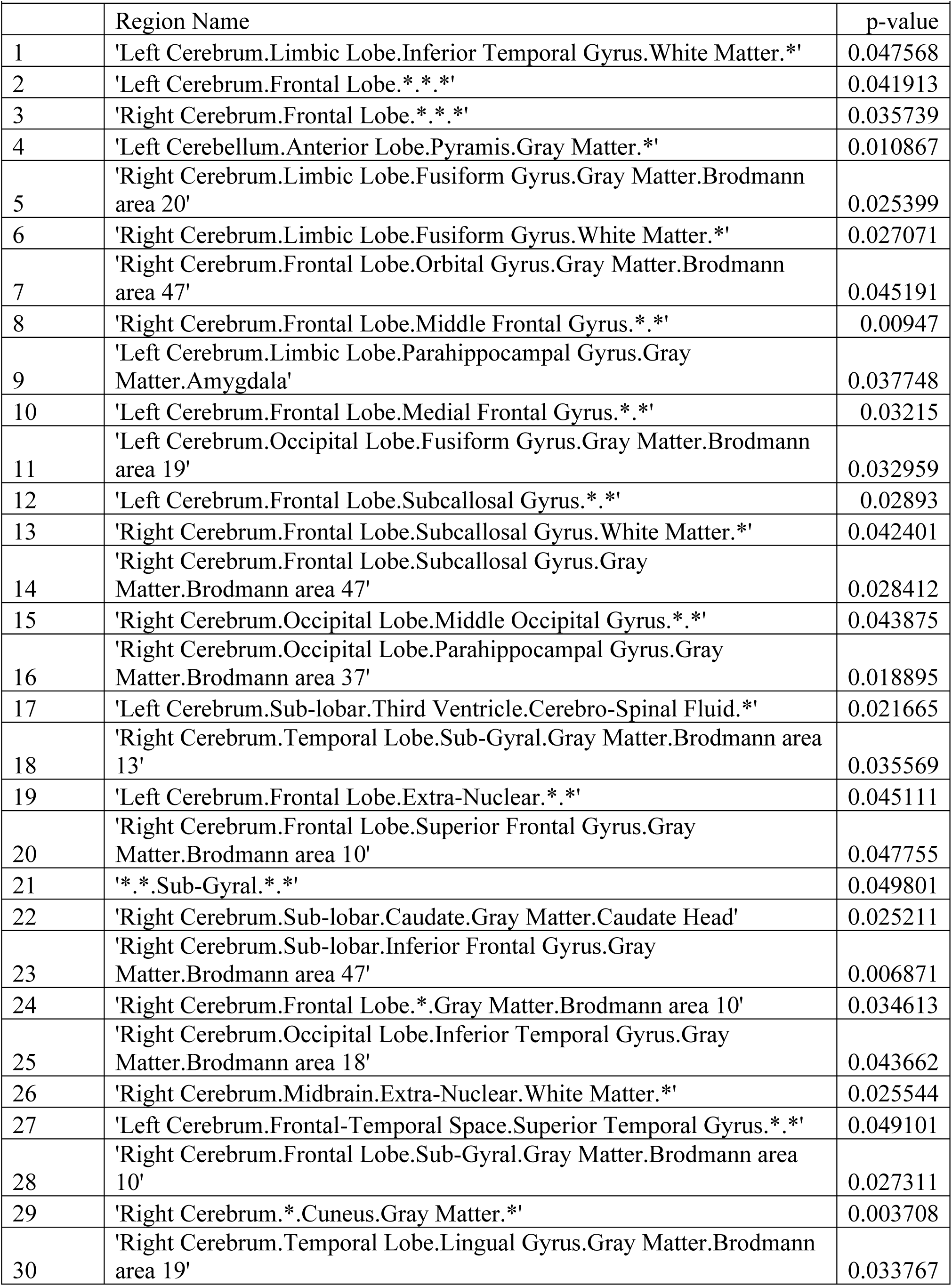
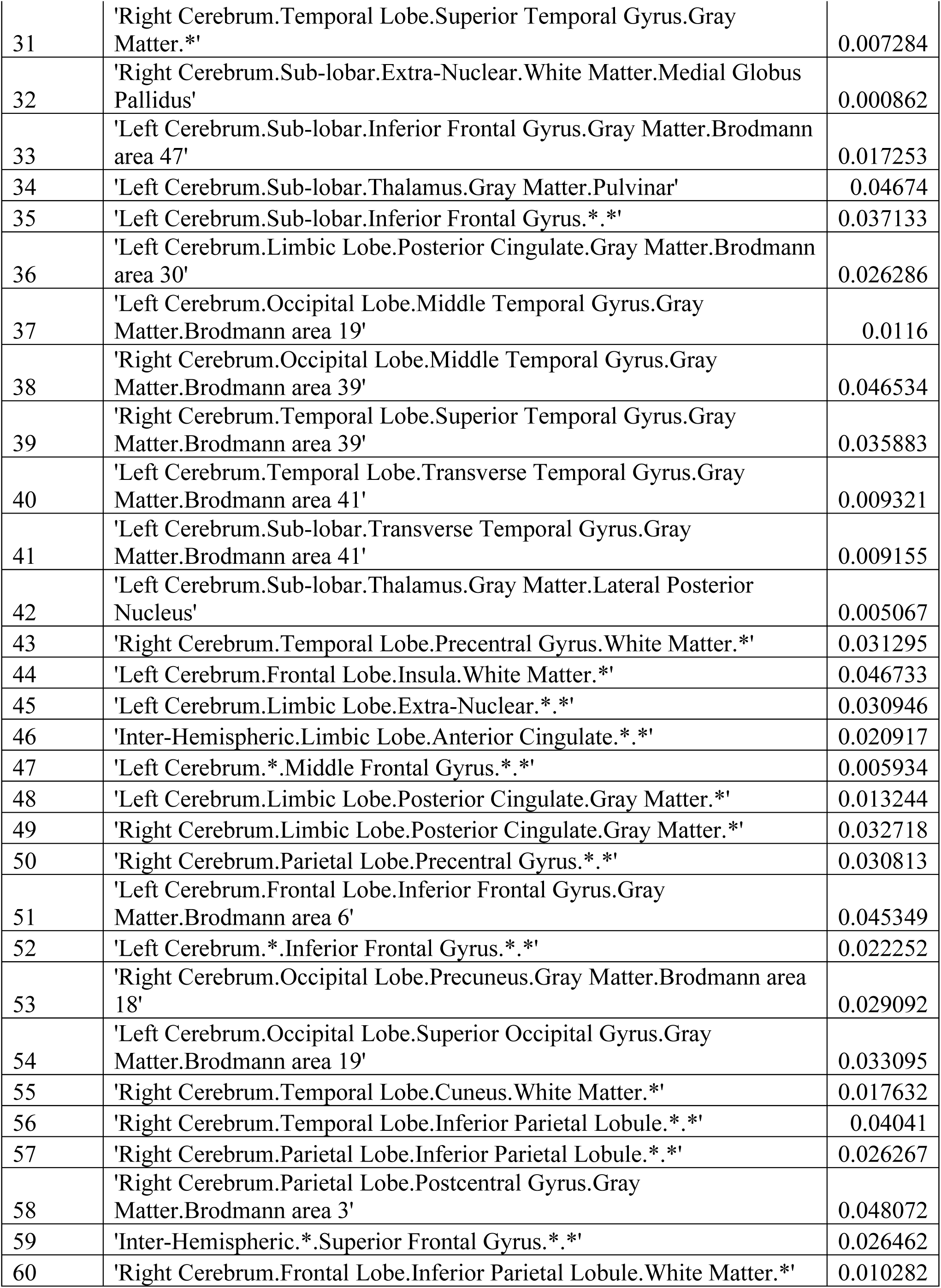
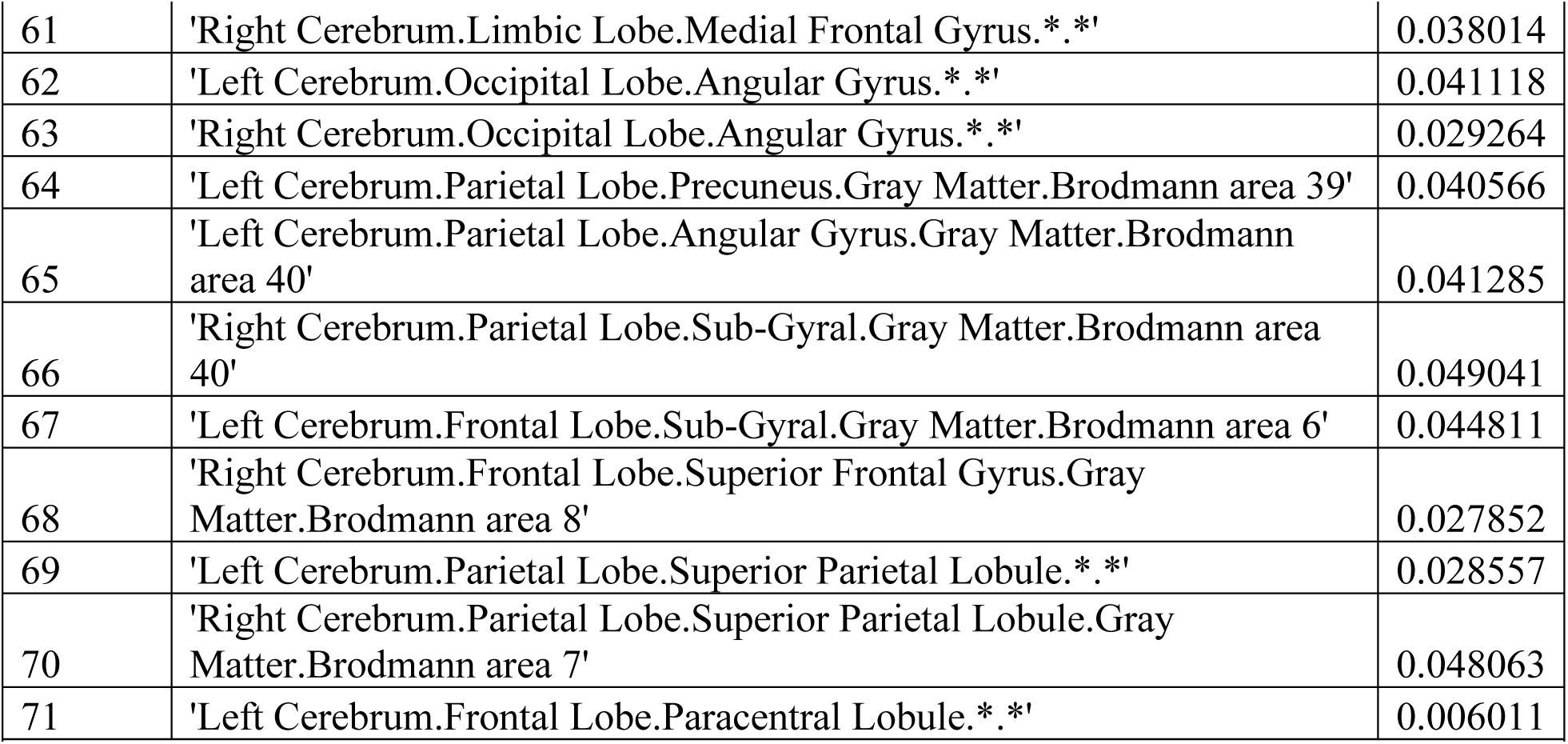
Significances between brains with CDR of 0 (n = 51) and 0.5 (n = 11).

**Table 2:**
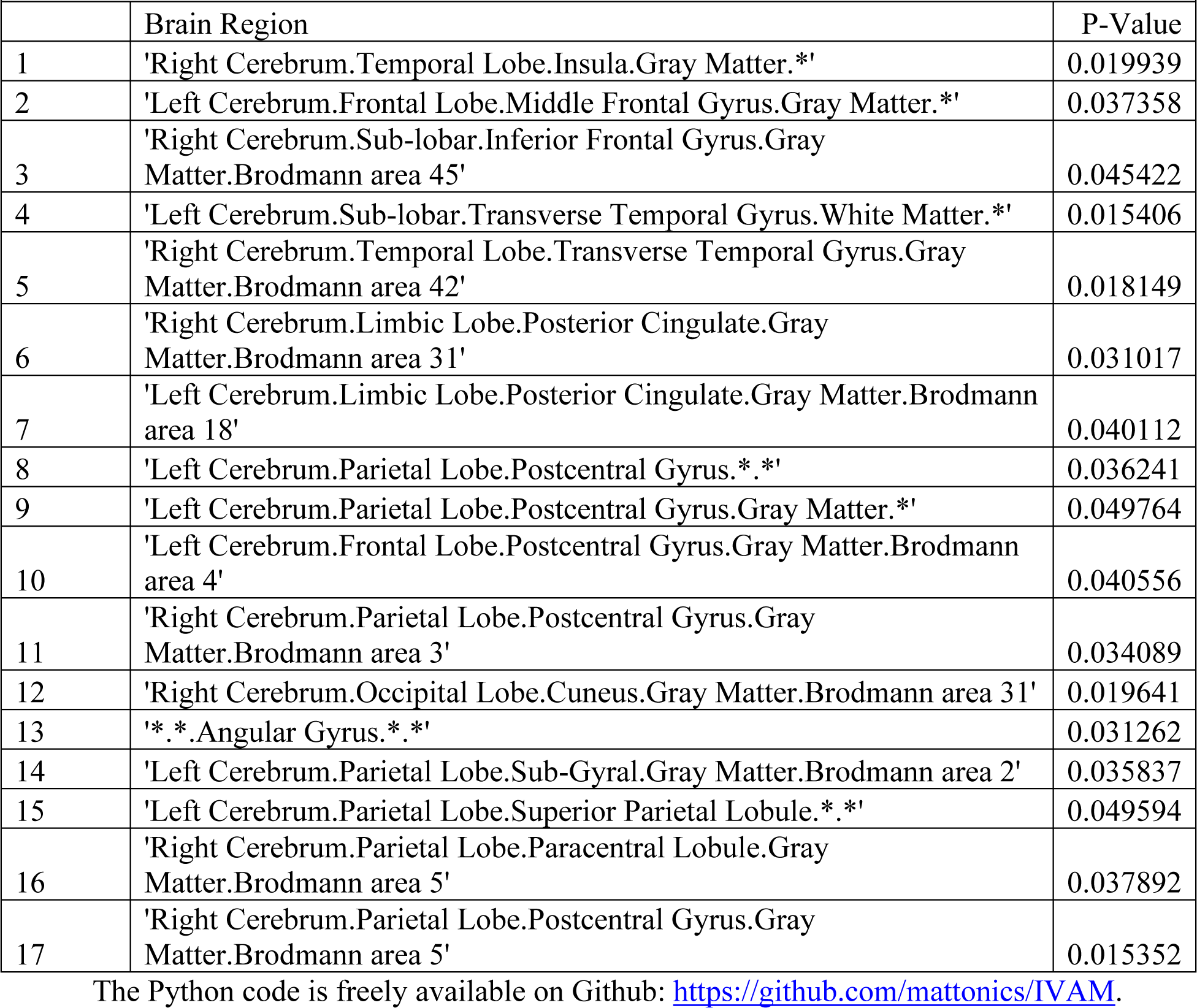
Significances between brains with CDR of 0 (n = 24) and 1 (n = 8).

## Conclusion

We created the best performing algorithm to-date for classifying 3D images taken of patients of neurodegenerative and neuropsychiatric diseases. In addition, the accuracy for voxel-by-voxel classification hovers around 83% as we labeled each voxel with the CDR of the brain. These diseases and disorders exhibit classification through many types of benchmarks that rate their severity. The IVAM code can be easily modified to accommodate different rating scales.

## Discussion

The skull-stripping algorithm we developed performs extremely well visually as shown for a single MRI image (Figure 2) separating the cortices from the skull as well as the cerebellum and lower brain from the skull in most instances. It performed successfully on a lot of the 380 images. There are examples of a missing lower brain, but nevertheless the algorithm achieved up to 90.0% classification accuracy. Adjustment of the dimensions of the structuring element could improve the results as well as a more robust thresholding algorithm such as a modified IsoData algorithm. A previously running version of IVAM which ran on spatially normalized images performed by OASIS authors but not skull-stripped by us achieved 92.2%. These data were pre-processed as a non-linear warping to the MNI152 template by OASIS authors.

**Figure 2:**
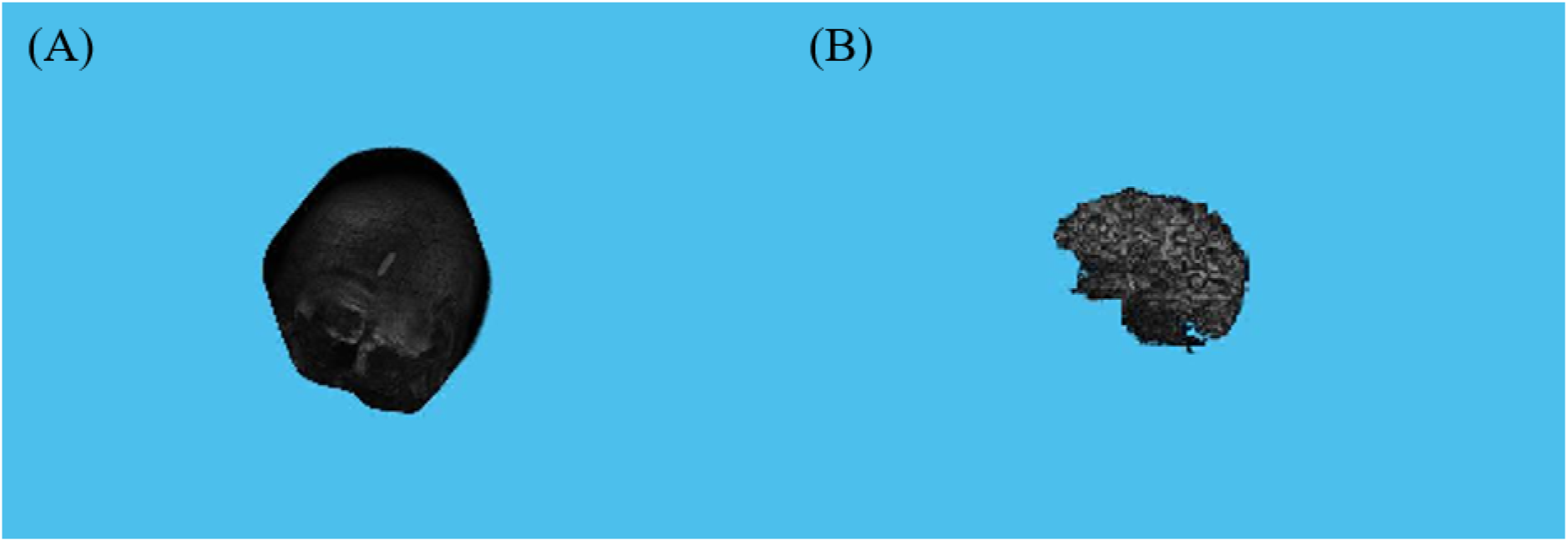
(A) Left panel illustrates the raw rendering of the 3D image obtained from OASIS. Right panel illustrates a brain with its skull image removed leaving behind gray and white matter in the image.

**Figure 3:**
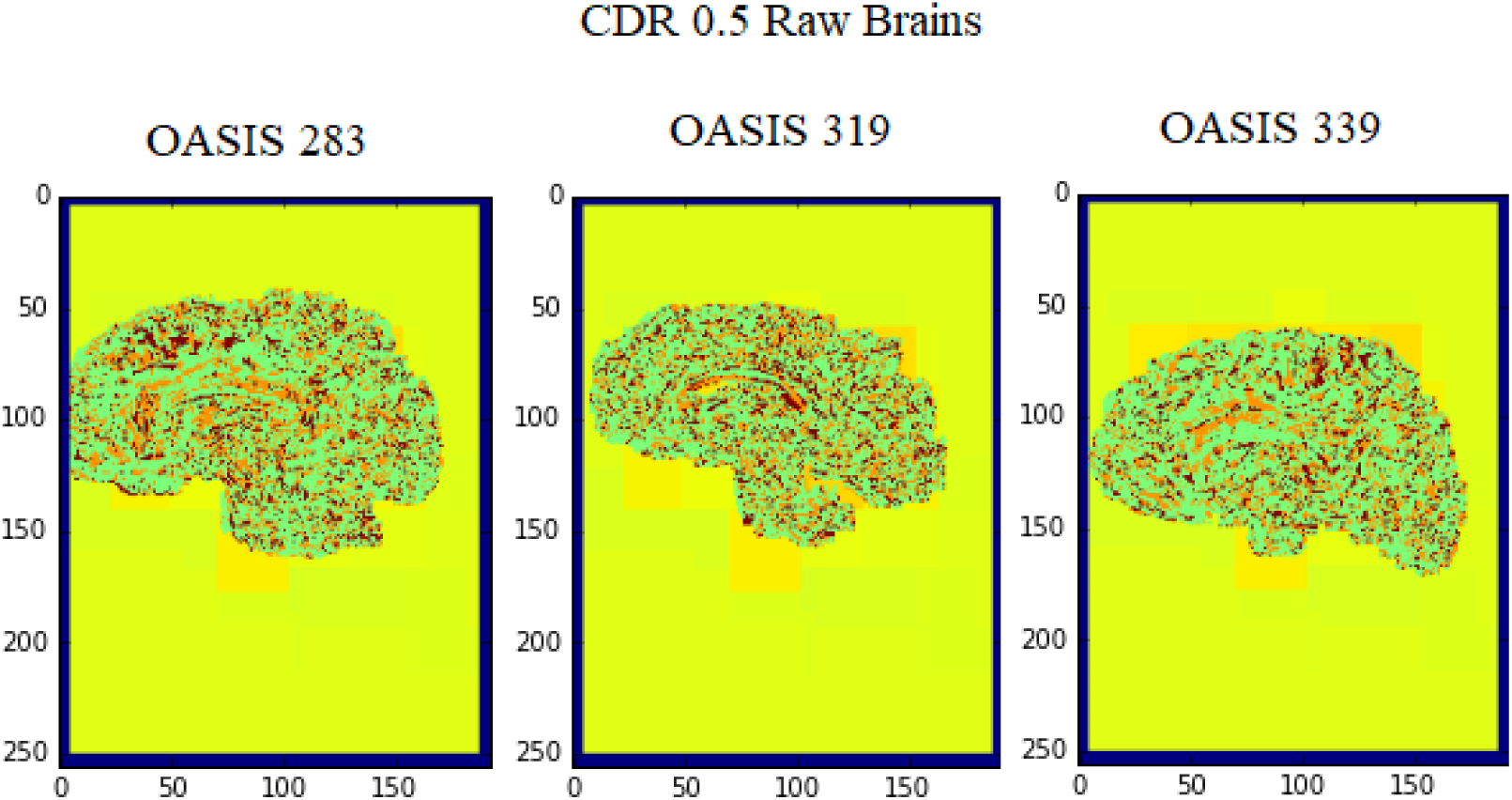
Shown are three sagittal cross-sections of brains afflicted by Alzheimer’s Disease with a CDR of 0.5. Two brains show high severity near the hippocampus while all three show moderate to severe severity near the inferior sagittal sinus. OASIS 339 illustrates how many brains afflicted by Alzheimer’s Disease show severity near the cortices separating brain lobes.

After running IVAM on nearly 200 images in the OASIS dataset, we find that a single extra-randomized tree regressor predicts with 90.0% accuracy when trained on around 90% of the dataset and tested on around 10% of the dataset, which represents the highest accuracy reported to-date on structural MRI images of Alzheimer’s Disease patients, as a tertiary regression on CDRs of 0, 0.5, and 1 in early AD and 83.3% accuracy as a binary regression of 0 and any higher CDR. We also find that the extra-randomized trees regressor as a forest (a bag of trees) predicts with 89.7% accuracy as a tertiary regression and 83.8% accuracy as a binary regression. We found that the single extra-randomized regressor tree predicts better than a bag of them. Current state-of-the-art prediction methods include a maximum of around 85% in binary classification as explained by Moscoso *et al*. 2019.

Next steps include improving the KNN part of the algorithm to accept non-integer values, i.e. CDR of 0.5, which should theoretically yield higher accuracy than the current tertiary classification, and improving identification of severely affected areas to a probabilistic map using the labeled MNI152 atlas which can be retrieved by installing MindBoggle.

## Data Availability

The code for IVAM is freely available on GitHub.

## Acknowledgements

We would like to thank Dr. Nigel S. Bamford for giving critiques on the performance of our algorithm and reviewing our writing.

